# The dark side of SARS-CoV-2 rapid antigen testing: screening asymptomatic patients

**DOI:** 10.1101/2021.04.24.21256040

**Authors:** Giorgia Caruana, Laure-Line Lebrun, Oriane Aebischer, Onya Opota, Luis Urbano, Mikael de Rham, Oscar Marchetti, Gilbert Greub

## Abstract

Most of the reports describing SARS-CoV-2 rapid antigen tests (RATs) performances derive from COVID-19 symptomatic subjects in outpatient settings during periods of highest incidence of infections and high rates of hospital admissions. Here we investigated the role of RATs in an Emergency Department, as a screening tool before admission for COVID-19 asymptomatic patients. Each patient was screened with two simultaneous nasopharyngeal swabs: one immediately analyzed at the bedside using RAT and the other sent to the laboratory for RT-PCR analysis. A total of 116 patients were screened at hospital admission in a 250-bed community hospital in Morges (EHC), Switzerland. With a disease prevalence of 6% based on RT-PCR results, RAT detected only two out of seven RT-PCR positive patients (sensitivity 28.6%) and delivered two false positive results (specificity 98.2%), thus resulting not fiable enough to be used as a screening method in this clinical scenario.

## 1. Introduction

The world has been dealing with SARS-CoV-2 pandemic since the first cases of pneumonia of unknown origin described in December 2019 in Wuhan, China ^1^. One year later, huge steps have been made in clinical knowledge on this new infectious disease and different types of diagnostic tests have been developed ^2^. Reverse transcriptase polymerase chain reaction (RT-PCR), which detects the viral RNA, is now largely considered as the diagnostic gold standard. However, questions remain regarding the optimal clinical use and the indications of antigen tests.

In Switzerland, the first epidemic wave (March-April 2020) forced laboratories to use the maximum of their test capacities. Because of the high flow of patients consulting hospitals’ Emergency Departments, faster diagnostic results were needed for triage, aimed at minimizing nosocomial transmissions. Rapid molecular systems detecting viral RNA, such as GeneXpert SARS-CoV-2 PCR test (Cepheid, USA), combined with classic RT-PCR systems, adequately responded to this clinical need, nevertheless with difficulties in reagents supplies.

In October 2020 Switzerland faced a massive second wave, with up to 1800 infections in 14 days/100.000 inhabitants ^3^, representing one of the world’s highest rate at that period. To meet the urgent need for rapid diagnosis immediately followed by quarantine and contact tracing, a key tool for optimal management of epidemics, the Swiss Federal Office of Public Health (FOPH) authorized the use of rapid antigen tests (RATs) in addition to gold-standard RT-PCR ^4^.

Several reports describe RAT performances, most of them derived from COVID-19 symptomatic subjects in outpatient settings during periods of highest incidence of infections and high rates of hospital admissions ^5,6^. To date, more data are needed to clarify the role of RAT as a screening tool among patients admitted to hospital Emergency Departments, with or without symptoms of COVID-19, during different phases of the epidemic curve. An extensive evaluation was performed in an Emergency Department of an Italian hospital with Standard Q ^®^ COVID-19 Rapid Antigen Test (SD Biosensor, Roche) screening on nasopharyngeal samples from symptomatic and asymptomatic patients ^7^. Among patients without COVID-19 symptoms, a sensitivity of 50% and a specificity of 99.6% was reported, with a SARS-CoV-2 prevalence of 6.5%. Another study performed in Germany investigated the use of RAT as a screening tool among symptomatic patients with COVID-19 admitted to the Emergency Department ^8^. In this study, performed with Standard Q ^®^ COVID-19 Rapid Antigen Test (SD Biosensor/Roche) on nasopharyngeal samples, sensitivity was 75.3% and specificity 100%, with a COVID-19 prevalence of 32.8%. After the implementation of RAT at the Emergency Department of the Lausanne University Hospital (CHUV) for quick triaging of patients with or without COVID-19 symptoms ^9^, we applied a combined RAT and RT-PCR nasopharyngeal screening to asymptomatic patients admitted in the Emergency Department of a 250-bed community hospital (EHC) in Morges, Switzerland.

## 2. Methods

Two simultaneous nasopharyngeal swabs were collected to screen asymptomatic adults hospitalized in medical and surgical wards according to a standard procedure of the Emergency Department of EHC. The first swab was analyzed at the bedside using Standard Q^®^ COVID-19 Rapid Antigen Test (SD Biosensor-Republic of Korea/Roche-Switzerland). Patients with a positive RAT were isolated in single rooms waiting for the molecular confirmation, performed on one of the following platforms at the Institute of Microbiology of the Lausanne University Hospital (CHUV): i) Test Cobas 6800® SARS-CoV-2 (Roche, Basel, Switzerland) or ii) automated high-throughput molecular diagnostic platform, using Magnapure RNA-extraction coupled to applied Biosystems 7900 amplification device (Quant Studio 7) and three Hamilton robots (with primers targeting the E- and RdRp-encoding) ^10^. RAT and RT-PCR results were compared in the frame of the EHC Patients Safety Program. This analysis in the frame of the CHUV Microbiology Laboratory Quality Control Program was approved by the institutional Ethical Review Board.

### Ethical declaration

This article was prepared according to STANDARD guidelines for diagnostic accuracy studies reporting. The data on the fiability of the different antigen assays were obtained during a quality enhancement project at our institution (CHUV, Lausanne). According to national law (Swiss Federal Act on Human Research), the performance and publishing the results of such a project can be done without asking the permission of the competent research ethics committee

## 3. Results

From 04/12/2020 to 04/01/2021, we consecutively screened 116 asymptomatic patients. 63 (54.3%) females and 53 (45.7%) males were tested, with a median age of 46.7 years [IQR 35.3-69.6] (population characteristics are described in Table 1).

**Table 1.** Characteristics of hospitalized patients without CoVID-19 symptoms admitted to the EHC Emergency Department between 4/12/2020 and 04/01/2021.

|  | Patients without CoVID-19 symptoms<br>(n=116) |  |
| --- | --- | --- |
|  | SARS-CoV-2<br>positive RAT<br>(n=4) | SARS-CoV-2<br>negative RAT<br>(n=112) |
| **Gender** |  |  |
| Female | 2 (50%) | 61 (54.5%) |
| Male | 2 (50%) | 51 (45.5%) |
| **Age** |  |  |
| Median [IQR], years | 41.9 [30.7-61.2] | 46.7 [35.3-69.6] |
| **SARS-CoV-2 RT-PCR**<br>**result** |  |  |
| Negative | 2 (50%)<sup>a</sup> | 107 (95.5%) |
| Positive | 2 (50%) | 5 (4.5%) |
| **Viral load** |  |  |
| Median [IQR], cp/ml | 1.8e+08<br>[1.8e+08 - 1.8e+08]<sup>b</sup> | 1.9e+04<br>[6.2e+03 - 8e+05] |

RAT: rapid diagnostic testing. RT-PCR: reverse transcriptase polymerase chain reaction. IQR: interquartile range. Cp/ml: viral copies per milliliter. <sup>a</sup> Among 4 positive RAT results, 2 were false positive results, not confirmed by RT-PCR. In this setting, RAT detected less than one third (2/7, 28.6%) of asymptomatic hospitalized patients, who resulted positive by RT-PCR. <sup>b</sup> One out of 7 RT-PCR positive patients (also tested positive with RAT) had a viral load at the limit of detection, that was not-quantifiable.

As compared to RT-PCR, RAT detected two out of seven SARS-CoV-2 positive patients and delivered two false-positive results, exhibiting a sensitivity of 28.6% and a specificity of 98.2%. The prevalence of SARS-CoV-2 carriage of 6% according to RT-PCR results was significantly underestimated by RAT to 1.7%.

## 4. Discussion

RATs are an attractive option for COVID-19 diagnostics due to low costs, rapidity and point-of-care solutions. However, they show a gap in analytical sensitivity as compared to the gold-standard RT-PCR, the detection of the viral load being reduced by a factor 1.000 to 10.000.

In Switzerland, RATs are authorized for immediate COVID-19 diagnosis in outpatients with symptoms lasting less than 4 days and early cohorting in-patients due to high numbers of hospitalizations, when pre-test probability is above 20% ^4^. This second condition was recommended because in settings with disease prevalence above 20%, the diagnostic performance gap might be partially compensated by the diagnostic speed, allowing prompt isolation of highly contagious patients, thus reducing the risk of nosocomial transmission.

Turcato and colleagues interestingly investigated the global clinical benefit derived from RAT screening against symptom-based screening in the Emergency Department with a decision curve analysis (DCA), reporting a considerable net benefit even in settings with SARS-CoV-2 prevalence lower than 15% ^7^. DCA is a useful, fast and easy alternative to a full decision analysis for giving a global overlook of benefits. Nevertheless, being only based on the reasonable range of threshold probabilities, DCA might have simplified some assumptions (e.g.: symptoms evaluation performed with the support of imaging/inflammatory parameters/clinical scores versus simple symptoms evaluation for initial triaging purposes) and more complex decision analyses might be needed before ultimately applying changes to screening strategy recommendations.

On the other hand, the data gathered so far showed a clinically relevant difference between RAT diagnostic performances in symptomatic and asymptomatic patients, with significantly lower sensitivity in the absence of symptoms. Hence, adopting a RAT-based screening strategy in patients without symptoms of COVID-19 might miss a significant number of asymptomatic SARS-CoV-2 carriers. This increases the risk of nosocomial transmission from patients with false-negative RAT results. Moreover, false-positive RAT results likely raise the hazard of hospital-acquired COVID-19, if the patient is cohorted with SARS-CoV-2 RT-PCR-positive patients.

These RATs limitations represent key issues for patients’ safety in the hospital arguing against the use of RAT screening among asymptomatic patients admitted via the Emergency Department.

## Data Availability

Data are not publicly available but they will be shared upon request for peer-review purposes.

## Data availability

Study data are not publically available but they will be shared upon request for the peer-review process.

## Patient Consent Statement

This article was prepared according to STANDARD guidelines for diagnostic accuracy studies reporting. The data on the viability of the different antigen assays were obtained during a quality enhancement project at our institution (CHUV, Lausanne, Switzerland). According to national law (Swiss Federal Act on Human Research), the performance and publishing of the results of such a project can be done without asking the permission of the competent research ethics committee and patients data were gathered as part of routine care.

